# Clinically relevant combined effect of polygenic background, rare pathogenic germline variants, and family history on colorectal cancer incidence

**DOI:** 10.1101/2022.01.20.22269585

**Authors:** Emadeldin Hassanin, Isabel Spier, Dheeraj R. Bobbili, Rana Aldisi, Hannah Klinkhammer, Friederike David, Nuria Dueñas, Robert Hüneburg, Claudia Perne, Joan Brunet, Gabriel Capella, Markus M. Nöthen, Andreas J. Forstner, Andreas Mayr, Peter Krawitz, Patrick May, Stefan Aretz, Carlo Maj

## Abstract

**Background and aims:** Summarised in polygenic risk scores (PRS), the effect of common, low penetrant genetic variants associated with colorectal cancer (CRC), can be used for risk stratification.

**Methods:** To assess the combined impact of the PRS and other main factors on CRC risk, 163,516 individuals from the UK Biobank were stratified as follows: 1. carriers status for germline pathogenic variants (PV) in CRC susceptibility genes (*APC, MLH1, MSH2, MSH6, PMS2)*, 2. low (<20%), intermediate (20-80%), or high PRS (>80%), and 3. family history (FH) of CRC. Multivariable logistic regression and Cox proportional hazards models were applied to compare odds ratios (OR) and to compute the lifetime incidence, respectively.

**Results:** Depending on the PRS, the CRC lifetime incidence for non-carriers ranges between 6% and 22%, compared to 40% and 74% for carriers. A suspicious FH is associated with a further increase of the cumulative incidence reaching 26% for non-carriers and 98% for carriers. In non-carriers without FH, but high PRS, the CRC risk is doubled, whereas a low PRS even in the context of a FH results in a decreased risk. The full model including PRS, carrier status, and FH improved the area under the curve (AUC) in risk prediction (0.704).

**Conclusion:** The findings demonstrate that CRC risks are strongly influenced by the PRS for both a sporadic and monogenic background. FH, PV, and common variants complementary contribute to CRC risk. The implementation of PRS in routine care will likely improve personalized risk stratification, which will in turn guide tailored preventive surveillance strategies in high, intermediate, and low risk groups.

## Introduction

Colorectal cancer (CRC) is the fourth leading cancer-related cause of death worldwide. Major established exogenous risk factors are summarized as Western lifestyle. However, an inherited disposition contributes significantly to the disease burden since up to 35% of interindividual variability in CRC risk has been attributed to genetic factors.^1,2^

Around 5% of CRC occur on the basis of a monogenic, Mendelian condition (hereditary CRC), in particular Lynch syndrome (LS) and various gastrointestinal polyposis syndromes. Here, predisposing rare, high-penetrance pathogenic variants (PV, constitutional / germline variants) result in a considerable cumulative lifetime risk of CRC and a syndrome-specific spectrum of extracolonic tumors. The autosomal dominant inherited LS is by far the most frequent type of hereditary CRC with an estimated carrier frequency in the general population of 1:300-1:500.^3–5^ It is caused by a heterozygous germline PV in either of the mismatch repair (MMR) genes *MLH1, MSH2, MSH6* or *PMS2* or, in few cases, by a large germline deletion of the *EPCAM* gene upstream of *MSH2*. The most frequent Mendelian polyposis syndrome is the autosomal dominant Familial Adenomatous Polyposis (FAP) caused by heterozygous germline PV in the tumor suppressor gene *APC*, followed by the autosomal recessive *MUTYH*-associated polyposis (MAP) which is based on biallelic germline PV of the base excision repair gene *MUTYH*.^6,7^ However, even in such monogenic conditions, the inter- and intrafamilial penetrance and phenotypic variability is striking, pointing to modifying exogenous or endogenous factors. Heterozygous (monoallelic) *MUTYH* germline PV may be associated with a slightly increased CRC risk (^8^;^9^); the carrier frequency in northern European populations is estimated to be 1:50-1:100.^3^

Approximately 20-30% of CRC cases are characterized by a suspicious, but unspecific familial clustering of CRC (familial CRC). Around 25% of CRC cases occur before 50 years of age (early-onset CRC); in around one quarter of those a hereditary type (mainly LS) has been identified.^10^ Although further high-penetrance candidate genes have been proposed, ^11–13^ the majority of familial and early-onset cases cannot be explained by monogenic subtypes and instead are supposed to result from a multifactorial / polygenic etiology including several moderate-/intermediate penetrance risk variants and shared environmental factors. A positive family history (FH) in first- and second-degree relatives increases the risk of developing CRC by 2-to 9-fold (^14^;^15^), which underpins the hypothesis of shared genetic and non-genetic risk factors.

A variety of models to predict CRC risk has been developed and evaluated, which include clinical data, FH, lifestyle factors, and genetic information.^16^ For more than a decade, genome-wide association studies (GWAS) in large unselected CRC cohorts identified an increasing number of common, low-penetrance risk variants, mainly single nucleotide polymorphisms (SNPs), which are significantly associated with CRC risk.^17–20^ Each SNP risk allele individually contributes only little to CRC risk (OR 1.05 to 1.5), however, summarised in quantitative polygenic risk scores (PRS), the combined effect might explain a substantial fraction of CRC risk variability and can identify individuals at several times lower and greater risk than the general population.^21–23^

As such, it is expected that the genetic background defined by the common risk variants may not only influence the occurrence of late-onset sporadic cases, but also modulate the risk of familial, early-onset, and hereditary CRC.^24^ Recent studies demonstrated that high PRS values are associated with an increased risk of CRC and other common cancers in the general population up to an order of magnitude that is almost similar to hereditary tumor syndromes.^25,26^

Based on these data, it can be hypothesized, that the identification of common genetic CRC risk variants not only provides deep insights into the biological mechanisms and pathways of tumorigenesis, but could improve personalized risk stratification for sporadic, familial / early-onset, and hereditary CRC in the future by the implementation of SNP-based PRS screening in routine patient care, which will in turn guide tailored preventive strategies in high, moderate, and low risk groups.

However, even if previous studies provide promising results for a clinical benefit of a PRS-based personalized risk stratification, the impact of common risk factors and their interplay with high-penetrance variants and other unspecified factors, captured partly by the FH, still has to be improved and validated in additional patient cohorts.

In the present work, we compare the prevalence and the lifetime risk of CRC among 163,516 individuals from a population-based European repository (UK Biobank, UKBB). Individuals were stratified according to three major risk factors 1) their carrier status of rare, high-penetrance pathogenic or likely pathogenic germline variants (hereafter defined as PV) in the MMR and the *APC* genes, 2) a low, intermediate, or high PRS, and 3) a FH for CRC.

## Material and methods

### Data Source

UK Biobank (UKBB) genetic and phenotypic data were used in this study. UKBB is a long-term prospective population-based study that has recruited volunteers mostly from England, Scotland, and Wales, with over 500,000 participants aged 40 to 69 years at the time of recruitment. For each participant, extensive phenotypic and health-related data is available; genotyping data is accessible for 487,410 samples, and exome sequencing data is available for 200,643 people. All participants gave written consent, and the dataset is available for research.^27^

### Study participants

CRC cases were defined based on self-reported code of 1022 or 1023 (in data field 20001), or ICD-10 code of C18.X or C20.X, D01.[0,1,2], D37.[4,5], or ICD-9 of 153.X or 154.[0,1] (in hospitalization records). Control samples were those that had no previous diagnosis of any cancer. The study includes people of all ethnicities. Outliers for heterozygosity or genotype missing rates, putative sex chromosome aneuploidy, and discordant reported sex versus genotypic sex were excluded. Only individuals (n=200,643) who had both genotyping and whole-exome sequencing (WES) data were considered. If the genetic relationship between individuals was closer than the second degree, defined as kinship coefficient > 0.0884 as computed by the UK Biobank, we removed one from each pair of related individuals (cases were retained if exist).

### Variant selection

We used ANNOVAR^28^ to annotate the VCF files from the 200,643 WES samples. The Genome Aggregation Database (gnomAD)^29^ were used to retrieve variant frequencies from the general population. We focused on rare PV for hereditary CRC (Lynch syndrome, polyposis) and considered the same variant filtering approach that was used in a recent study aiming at selecting rare PV.^30^ The following inclusion criteria were used: 1) only *APC, MUTYH, MLH1, MSH2, MSH6, PMS2* variants in protein-coding regions were included; 2) allele frequency (AF) <0.005 in at least one ethnic subpopulation of gnomAD; 3) not annotated as “synonymous,” “non-frameshift deletion” and “non-frameshift insertion”; 4) annotated as “pathogenic” or “likely pathogenic” based on ClinVar.^31^ We did not include *MUTYH* in the pooled analysis since no biallelic (i.e. high penetrance) case was identified in the cohort; however, we included the heterozygous (monoallelic) carriers in the single gene analysis to compare the effect size with the other genes.

### PRS

We applied a previously validated PRS for CRC with 95 variants to calculate the PRS.^17^ The PRS was computed using the PLINK 2.0^32^ scoring function through UKBB genotype data. To reduce PRS distributions variance among genetic ancestries, we applied a previous approach.^33^ We used the first four ancestry principal components (PCs) to fit a linear regression model to predict the PRS across the full dataset (pPRS∼PC1 + PC2 + PC3 + PC4). Adjusted PRS (aPRS) were calculated by subtracting pPRS from the raw PRS and used for the subsequent analysis.

### Statistical analysis

Individuals were divided into groups depending on 1) carrier status of PV, 2) PRS, and 3) FH. For FH, we considered participants’ reports of CRC in their parents and siblings (data fields: 20110, 20107, 20111). For PRS, individuals were assigned into three groups: low (<20% PRS), intermediate (20-80% PRS), and high (>80% PRS).

We conducted both an analysis specific to single genes and a combined analysis (i.e., carriers of PV in *APC, MLH1, MSH2, MSH6* and *PMS2*). First, we estimated the OR for each carrier group based on a logistic regression adjusting for age at recruitment, sex and the first four ancestry PCs. Afterwards, we additionally incorporated interactions between PV carriers and FH with PRS by introducing an interaction term within the logistic regression model.

We calculated the lifetime risk by age 75 from carrier status of rare PV and the PRS based on a Cox proportional hazards model. Individual’s age served as the time scale, representing the time to event, for observed cases (age at diagnosis), and censored controls (age at last visit). Carrier status, PRS category, age, sex and the first four ancestry PCs were incorporated in the model, and adjusted survival curves were produced.

Model performance was assessed via the area under the receiver operating characteristic curve (AUC), Nagelkerke’s Pseudo-R^2^, and the C-index for time-to-event data. R 3.6.3 with the corresponding add-on packages *survival* and *survminer* was used for all statistical analyses.

## Results

### Stratification of UKBB individuals for CRC prevalence, FH, and PV carrier status

We identified 1,902 CRC cases (894 prevalent cases and 1,008 incident cases) among the 163,516 UKBB individuals that retained after exclusion criteria, with a mean age at diagnosis of 60.9 years. The remaining 161,614 individuals with no previous diagnosis of any cancer were considered as controls, with a mean age of 56.9 years at last visit (Table 1). The European population represents 92% of the analyzed cohort.

**Table 1:** Characteristics of the 163,516 UK Biobank participants by colorectal cancer (CRC) status.

|  | Cases | Controls |
| --- | --- | --- |
| Participants, n | 1902 | 161614 |
| Male, n (%) | 1017 (53.47) | 73979 (45.78) |
| Female, n (%) | 885 (46.53) | 87635 (54.22) |
| Age, mean (SD) | 60.96 (8.56) | 56.91 (8.51) |
| Carriers, n (%) | 30 (1.58) | 369 (0.23) |
| Family history of colorectal cancer, n (%) | 368 (19.35) | 17696 (10.95) |

The fraction of individuals with a positive FH of CRC is significantly higher in cases (19%) compared to controls (11%) (OR = 1.95 [1.73-2.19], P < 0.01) and ranges between 9% and 23% in the subgroups (Table 2). There is a significantly higher proportion of individuals with a FH of CRC not only among carriers of PV in the selected cancer susceptibility genes (OR = 1.96 [1.72-2.20], P < 0.01), but also among non-carriers with high PRS (OR = 1.60 [1.31-1.94], P < 0.01).

**Table 2:**
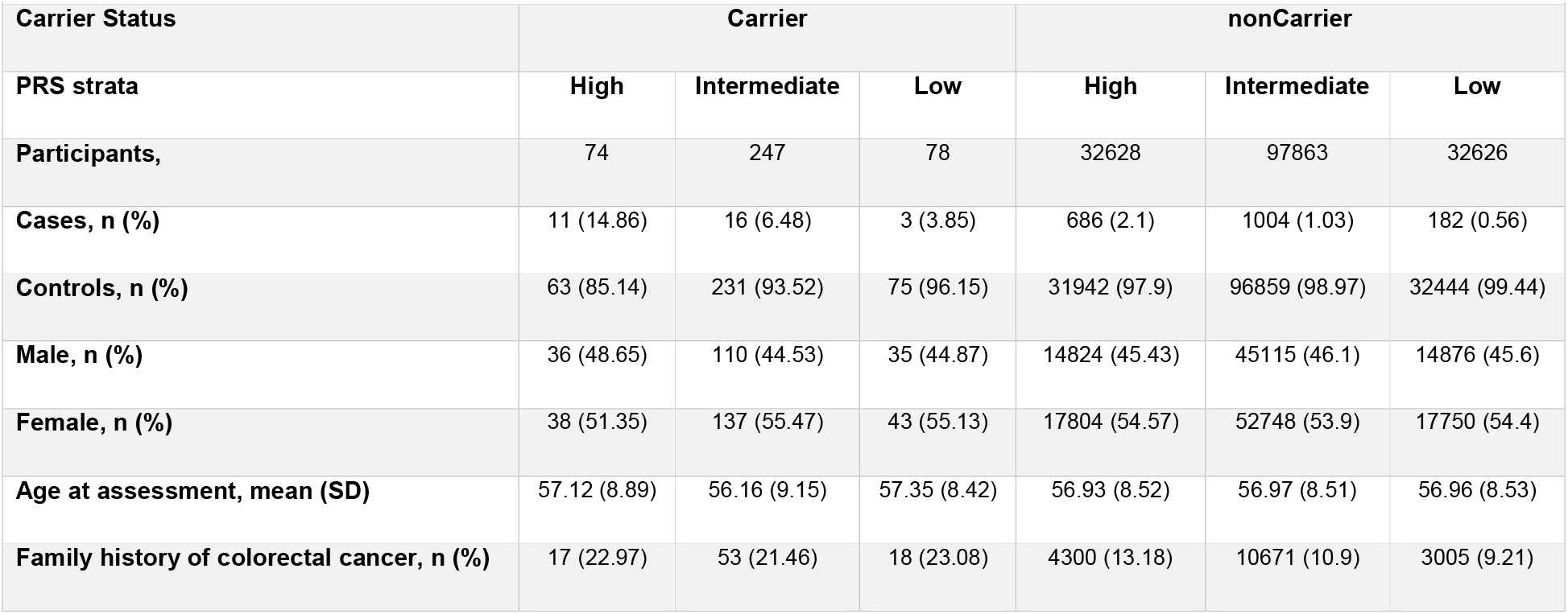
Characteristics of the UK Biobank participants by carrier status and polygenic risk score (PRS) strata.

| Carrier Status | Carrier |  |  | nonCarrier |  |  |
| --- | --- | --- | --- | --- | --- | --- |
| PRS strata | High | Intermediate | Low | High | Intermediate | Low |
| Participants, | 74 | 247 | 78 | 32628 | 97863 | 32626 |
| Cases, n (%) | 11 (14.86) | 16 (6.48) | 3 (3.85) | 686 (2.1) | 1004 (1.03) | 182 (0.56) |
| Controls, n (%) | 63 (85.14) | 231 (93.52) | 75 (96.15) | 31942 (97.9) | 96859 (98.97) | 32444 (99.44) |
| Male, n (%) | 36 (48.65) | 110 (44.53) | 35 (44.87) | 14824 (45.43) | 45115 (46.1) | 14876 (45.6) |
| Female, n (%) | 38 (51.35) | 137 (55.47) | 43 (55.13) | 17804 (54.57) | 52748 (53.9) | 17750 (54.4) |
| Age at assessment, mean (SD) | 57.12 (8.89) | 56.16 (9.15) | 57.35 (8.42) | 56.93 (8.52) | 56.97 (8.51) | 56.96 (8.53) |
| Family history of colorectal cancer, n (%) | 17 (22.97) | 53 (21.46) | 18 (23.08) | 4300 (13.18) | 10671 (10.9) | 3005 (9.21) |

In the analyzed CRC susceptibility genes *APC, MLH1, MSH2, MSH6, PMS2*, we identified 399 heterozygous carriers of 111 PV. They were present in 30 (1.57%) cases and 369 (0.23%) controls, which is in line with published data. A list of the considered variants and annotations is shown in Supplementary Table S1, a summary of the number of PV carriers per gene is provided in Supplementary Table S2. No individual with a homozygous PV was identified.

### PRS distribution within the UKBB cohort

CRC PRS follow a normal distribution both regarding raw and PC-adjusted PRS (Supplemental Figure S1) and is significantly higher in cases compared to controls (P < 2.2e™16) (Supplemental Figure S2).

The prevalence of CRC according to PRS percentiles demonstrates that values in the extreme right tail of the PRS distribution are associated with a non-linear increase of CRC risk, whereas in the left tail a less evident non-linear decrease can be observed (Supplemental Figure S3). This supports the hypothesis of using PRS to stratify individuals into risk classes (i.e., low, intermediate, and high risk) according to a liability threshold model.

### Interplay between PV and PRS

There was no overlap between the selected rare high penetrance PV and the common SNPs used for PRS calculation, and thus, the PRS represents an additional genetic signal. Notably, the PRS distributions showed that the mean and median of PRS is significantly higher in affected carriers compared to unaffected carriers (*P* = 0.004) (Supplemental Figure S4).

We assessed how CRC risk is influenced by PRS and carrier status for PV in high penetrant CRC susceptibility genes (*APC, MLH1, MSH2, MSH6, PMS2*) by calculating the ORs for CRC across groups compared to non-carriers with intermediate PRS as reference group. Non-carriers with a low or high PRS are estimated to have a 0.5-fold or 2.1-fold change in the odds for CRC, respectively. We observed that the PRS also alters the penetrance of PV in susceptibility genes considerably as PV carriers with high PRS had four times higher OR than carriers with low PRS (OR = 17.5 and 3.9, respectively; Figure 1A). We did not observe a significant interaction between PV carrier status and PRS (*p* = 0.87).

**Figure 1:**
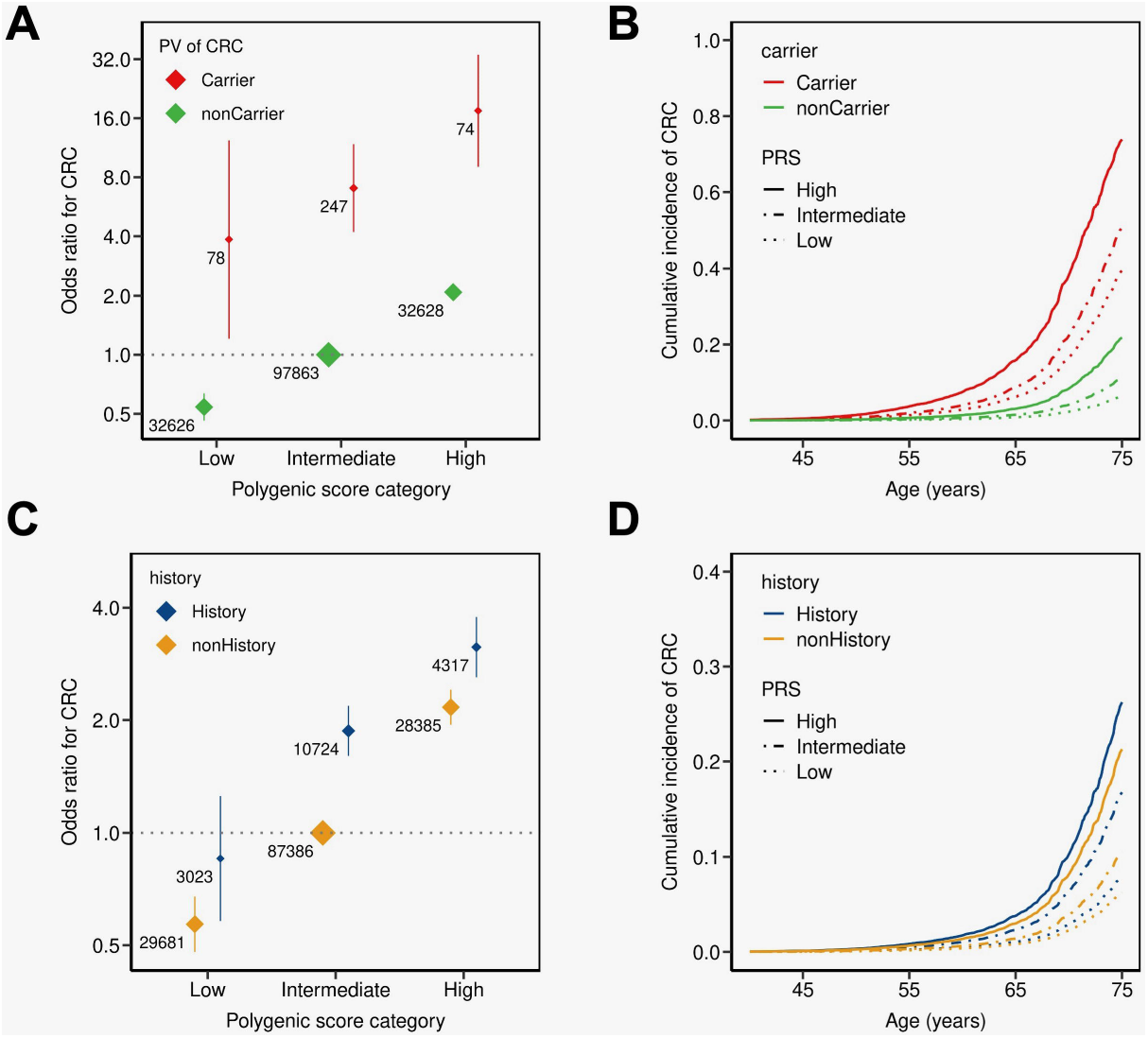
Colorectal cancer (CRC) odds ratio (OR) and cumulative incidence (CI) among individuals stratified for presence of pathogenic variant (PV) carrier status and family history. Individuals stratified for PV carrier status (A+B), and family history (first-degree relative with CRC) (C+D) into three strata based on their polygenic risk score (PRS): Low (<20 % percentile), intermediate (20-80 % percentile), or high (>80 % percentile) PRS. The OR was calculated from a logistic regression model with age, sex and the first four principal components of ancestry as covariates. The reference group was non-carriers with intermediate PRS (A), and no family history with intermediate PRS (C). The adjusted OR is indicated by the colored boxes. The numbers next to the ORs indicate the sample size of the corresponding group. The 95% confidence intervals are indicated by the vertical lines around the boxes. Cumulative incidence was estimated from a cox-proportional hazard model using age, sex and the first four ancestry principal components as covariates.

The high PRS, which is by definition present in 20% of the non-carriers, is associated with an almost doubled CRC risk (Figure 1A, Table 2). Since the vast majority (97.9%) of non-carriers are controls (=healthy), almost the same percentage results if only healthy non-carriers are considered.

Similarly, the lifetime cancer risk analysis shows a combined impact of PV and PRS: Among carriers, the estimated cumulative incidence by age 75 increased from 40% in case of a low PRS to 74% in case of a high PRS compared to 6% to 22% for non-carriers (Figure 1B).

### Inclusion of family history on cancer risk stratification

Taking individuals with no FH and intermediate PRS as a reference, both FH and PRS are associated with a higher CRC risk (Figure 1C). The CRC risk for individuals having low PRS and no FH (OR 0.6) is five times lower than for individuals having both positive FH and high PRS (OR 3.1). We did not observe a significant interaction between FH status and PRS (*p* = 0.12). Noteworthy, individuals without FH and high PRS and individuals with FH and intermediate PRS both have similar CRC risks with an OR of around 2, whereas the CRC risk of individuals having low PRS even in the context of a FH is decreased compared to the reference group.

Among individuals with FH, the cumulative CRC incidence by age 75 increases 3-fold from 8% in case of a low PRS to 26% in case of a high PRS (Figure 1D). Noteworthy, the cumulative CRC incidence of individuals with a positive FH and an intermediate PRS is lower (16%) than for individuals with negative FH and a higher PRS category (21%), respectively.

The full model integrating PRS, FH, and PV status shows that the CRC risk is strongly influenced by PRS in all groups (Figure 2, Supplementary Table S3). Considering the non-carriers with no FH and intermediate PRS group as reference, the CRC OR in low PRS is 0.6 for non-carriers with no FH, while it is estimated more than 60 times higher (OR 40) for carriers with FH and high PRS (Figure 2A). The corresponding cumulative CRC incidences are 6% and 98%, respectively (Figure 2B). Although all PV carriers showed a significantly increased CRC risk, both the PRS and FH modify these risks considerably: depending on the FH and PRS, the OR in PV carriers vary between 4 and 40 and the cumulative incidence between 35% and 98%.

**Figure 2:**
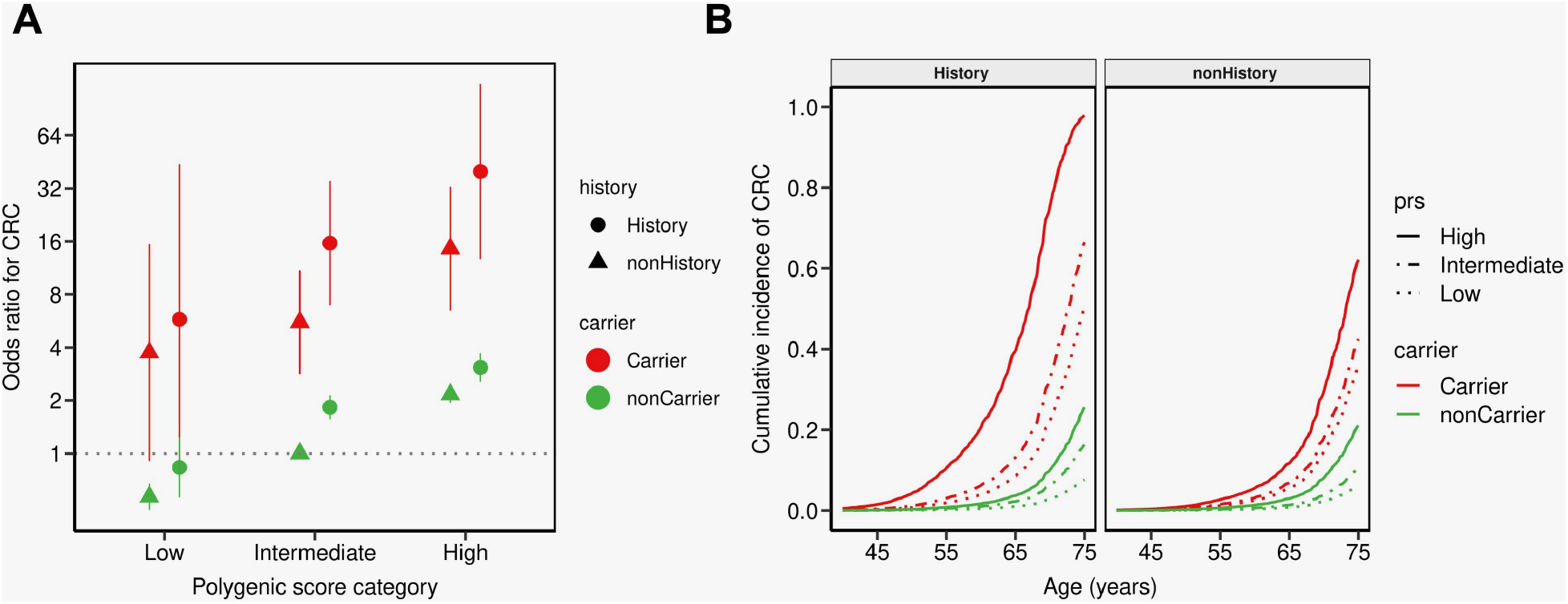
Interplay of pathogenic variant (PV) carrier status, family history (FH), and polygenic risk score (PRS). (A) Colorectal cancer (CRC) odds ratios (ORs) were estimated from logistic models adjusted for age, sex and first four ancestry principal components. Non-carriers with intermediate PRS and no family history served as the reference group. (B) Cumulative incidence was estimated from a cox-proportional hazard model using age, sex and the first four ancestry principal components as covariates.

PRS improved model discrimination over carrier status and FH of CRC in first-degree relatives. The AUC derived from PRS (0.688) was higher compared to those derived using FH (0.654) and carrier status (0.646). The full model including PRS, carrier status, and FH improved the AUC (0.704) in risk prediction by 1.6%, 5%, and 5.8%, respectively, and was also better than any combination of two factors (Table 3).

**Table 3:** Model discrimination assessed for combinations of polygenic risk score (PRS), family history of CRC (FH) and carrier

|  | **AUC (C.I 95%)** | **C-index** | **Nagelkerke's Pseudo-R<sup>2</sup>** |
| --- | --- | --- | --- |
| **PRS + FH + carrier** | 0.704 (0.68-0.73) | 0.657 | 0.055 |
| **PRS + FH** | 0.698 (0.67-0.72) | 0.652 | 0.052 |
| **PRS + carrier** | 0.693 (0.66-0.71) | 0.646 | 0.051 |
| **PRS** | 0.688 (0.66-0.71) | 0.640 | 0.049 |
| **FH + carrier** | 0.660 (0.64-0.69) | 0.580 | 0.032 |
| **FH** | 0.654 (0.63-0.68) | 0.574 | 0.031 |
| **Carrier** | 0.646 (0.62-0.67) | 0.556 | 0.030 |

Note: All models are adjusted for sex, age and first four ancestry principal components.

### The impact of polygenic risk in single gene mutation carriers

The gene-specific analysis revealed a strong variability in risk conferred by rare heterozygous PV in the different genes. The largest effect sizes are attributable for *MLH1* and *APC*, those for *MSH2* and *MSH6* are a bit less, while the effect size for *PMS2* is considerably lower (Figure 3). When heterozygous *MUTYH* variants are included in this analysis, the risks are very similar to the *PMS2*-related risks. Both the *PMS2* and heterozygous *MUTYH* risks show a broad overlap with the non-carrier risks, while there is no overlap between the risks of non-carriers and those with PVs in *MLH1, MSH2, MSH6*, and *APC*.

**Figure 3:**
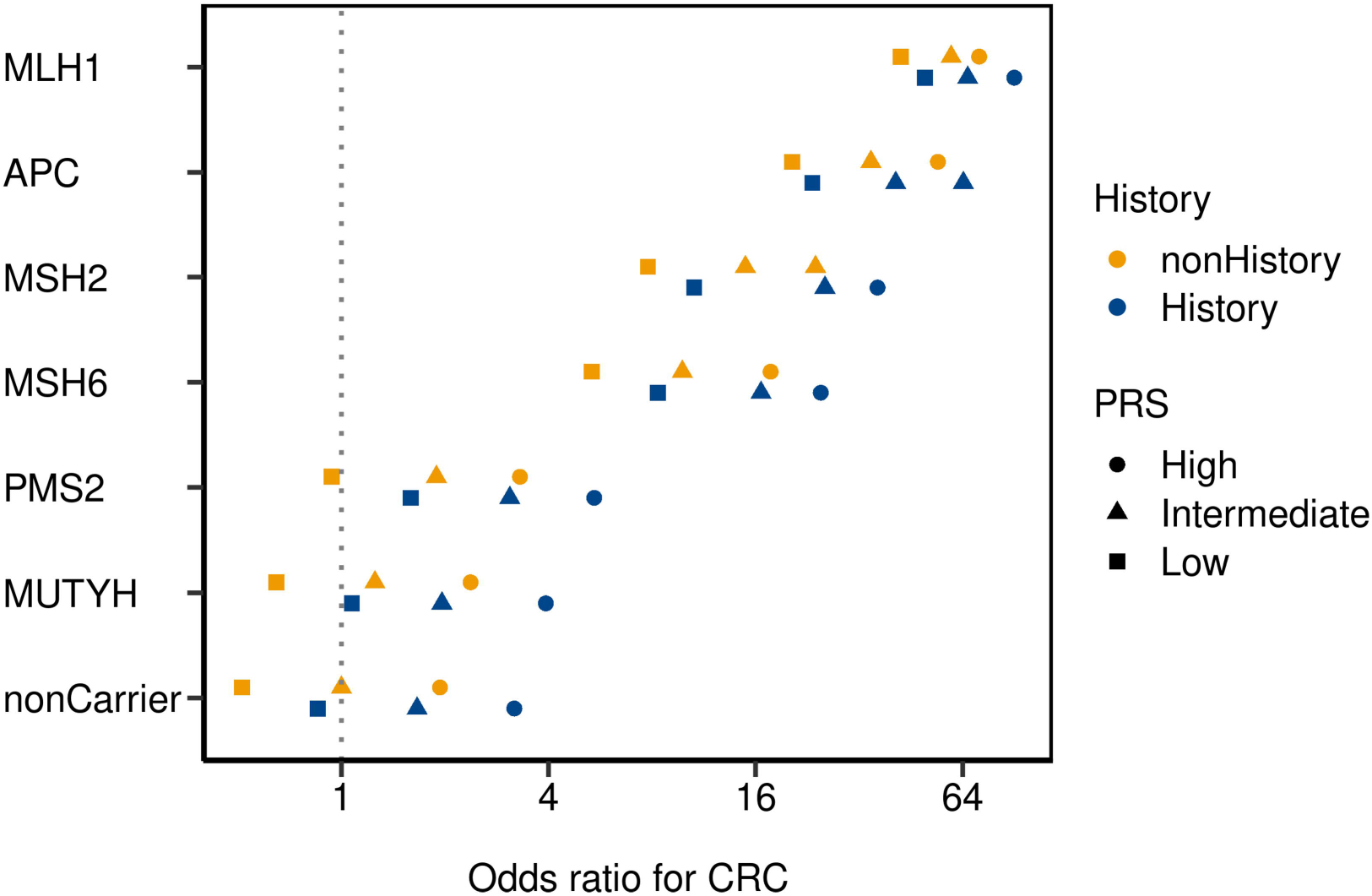
Interplay of heterozygous pathogenic variant (PV) carrier status, family history (FH), and polygenic risk score (PRS) in single genes. Odds ratios (ORs) for colorectal cancer (CRC) were estimated from logistic models adjusted for age, sex and first four ancestry principal components. Non-carriers with intermediate PRS and no family history served as the reference group.

We estimated how PRS and FH influence CRC prevalence among PV carriers in each of the five susceptibility genes. Despite the different effect sizes, the PRS and FH modifies the relative risk across all genes; however, the effect of PRS and FH is conversely related to the penetrance of the gene with the smallest effects in *MLH1* PV carriers.

As for the overall analysis, in the gene-specific analysis a positive FH, a PV in a cancer risk gene, and a high PRS are associated with an increased CRC risk. As such, an individual with a low-penetrance *PMS2* PV, but high PRS and / or positive FH ends up with an estimated CRC risk similar to a *MSH6* PV carrier without FH and / or low PRS.

## Discussion

Recent studies demonstrated that the polygenic background, defined as PRS based on disease-associated SNPs, modifies the risks for several cancers of the general population including CRC considerably, both in terms of age at onset and cumulative lifetime risks.^11,22,26,34–36^ In line with this, the risk alleles of those SNPs are found to also accumulate in unexplained familial and early-onset CRC cases.^24,37^ Whereas a low polygenic burden decreases the CRC risk down to one quarter on average, individuals with a high PRS (>80%) doubles and those with a very high PRS (99%) almost quadruplicate their risk and thus, reach a CRC risk in an order of magnitude almost comparable to carriers of hereditary CRC with low PRS.^30^ In a pervious study, Jia et al. found that the risk of CRC is significantly associated with its PRS: Compared with individuals in the lowest PRS quintile those in the highest quintile had a greater than threefold risk (during a 5.8-year follow-up period). Hazard Ratios estimated with the middle quintile as the reference resulted in a risk between 0.56-1.71, a threefold risk in those in the top 1% of PRS, and a 70% reduced CRC risk for individuals in the bottom 1% of the PRS.^36^

To extend these studies on how the CRC prevalence is influenced by genetic susceptibility using, we used the sufficiently larger, more robust dataset of the most recent UKBB cohort, incorporate the family history (FH) as an additional factor for risk stratification, and include a single gene analysis. We considered both the genetic component driven by rare high-penetrance PV associated with hereditary CRC and common low-penetrance variants captured by the PRS.

Firstly, our results confirm that the polygenic background strongly modulates CRC risk in the general population. Compared to the average polygenic burden, individuals with a low (<20%) or high (>80%) PRS are estimated to have a 0.5-fold or 2.1-fold change in the odds for CRC, respectively. The additional time-to-event analysis revealed a corresponding cumulative lifetime risk of 6% and 22% by age 75. Hence, when the PRS is included in risk calculation, around 20% of healthy individuals of the general population with no FH of CRC have a doubled CRC risk, which is similar to those with a first degree relative affected by CRC.^38^ These so far unknown and otherwise unrecognisable at-risk individuals might need surveillance 10-15 years earlier than usually recommended.^39^ On the other hand, the around 20% of individuals with low PRS and no FH might need less surveillance than the general population due to a considerably lowered risk, while even those with low PRS and positive FH might not need a more intense surveillance than the general population..

It is well known that among patients with hereditary CRC syndromes, the age of onset and cumulative CRC incidence is very heterogeneous, even within PV carriers of the same family. The estimated gene-specific, individual CRC lifetime risks of LS patients with *MLH1* or *MSH2* PV can be lower than 10% but as high as 90%-100% in a considerable fraction. In the past, the analysis of modifying effects based on common CRC-associated variants in LS and other high-risk groups has been restricted to selected cohorts and small subsets of SNPs.^40,41^ A recent study demonstrated that the polygenic background also substantially influences the CRC risk in LS using UKBB data, even though the ORs for CRC risks could only be predicted due to the small sample sizes.^30^ In the present work, ORs could be calculated directly from the model since over three times more UKBB individuals have been included with six times more CRC cases, and five times more PV carriers.

So secondly, we were able to show that the PRS modifies the CRC risks not only in the general population considerably, but also in carriers of a MMR gene PV identified in the general population. For the first time we demonstrated, that this is also true for *APC* PV. Depending on the PRS, the cumulative CRC lifetime incidence in PV carriers ranged between 40% and 74%, and thus, the PRS is able to explain parts of the interindividual variation in CRC risk among PV carriers.

However, the single-gene analysis revealed heterogeneous effects across genes and therefore the modifying role of the polygenic background should be framed within the absolute risk attributable to individual genes. As expected, the effect of the PRS seems to be relevant in particular in less penetrant CRC risk genes such as *PMS2* where the OR ranges between 0.94 and 5.43 respectively (Figure 3). This is in line with findings in moderate breast cancer risk genes such as *CHEK2, PALB2* and *ATM*^42–44^ and suggests that PRS inclusion in risk stratification may in particular be relevant to prevent excess of surveillance measures in PV carriers of those genes.

In addition, our results provide evidence that the inclusion of FH can further and independently improve the risk stratification in both carriers and non-carriers. Including PRS and FH in risk assessment, the cumulative CRC lifetime incidence ranged between 8% and 26%, and in PV carriers between 30% and 98%, and thus, outperformed the consideration of a single risk factor. This suggests that familial clustering points to additional risk factors besides those captured by common low-risk SNPs (PRS) and rare PV.^45,46^ These might be common and rare structural genetic alterations including copy number variants, rare non-coding variants, or other intermediate and low-impact risk variants not included routinely in PRS models, and non-genetic contributors such as environmental / lifestyle factors.

Only few PRS studies considered the FH. In line with our results, Jenkins et al. found no correlation between SNP-based and FH-based risks and an improved risk stratification when both PRS and FH are considered.^45^ In the analyses by Jia et al., the AUC derived from PRS (0.609) was substantially higher compared to the one derived using FH (0.523). Adding PRS and FH of cancer in first-degree relatives improved the models discriminatory performance (AUC 0.613).^16,47^ Our AUC calculations point in the same direction with a higher AUC (0.704) when all three risk factors (PRS, FH, carrier status) are considered.

Interestingly and in apparent contrast to our results and those of others, a study using 826 European-descent carriers of PV in the DNA MMR genes *MLH1, MSH2, MSH6, PMS2*, and *EPCAM* (i.e. LS carriers) from the Colon Cancer Family Registry (CCFR) did not find evidence of an association between the PRS and CRC risk, irrespective of sex or mutated gene, although an almost identical set of SNPs was used for PRS calculations.^48^ A reason which might partly explain different risk estimates between studies using individuals from a population-based repository such as the UKBB and those using curated clinical data registries, where patients / families with suspected hereditary disease are included (e.g. the CCFR), is a potentially different risk composition across cohorts recruited in different ways (recruitment bias). That way, a familial clustering of CRC might reflect the existence of several genetic and non-genetic risk factors as outlined above, which are not captured by the PRS and which may superimpose the polygenic impact.

In particular, the composition of cases and controls is different between the Jenkins et al.^48^ study on the one hand and the Fahed et al.^30^ and present study on the other hand. In the Jenkins et al. study, obviously both cases (i.e., PV carriers with CRC) and controls (healthy PV carriers) derived from the same LS families, while the UKBB controls are PV carriers not apparently related to the PV cases. This is also reflected by the different ratio between cases and controls (7.5% CRC cases among PV carriers in the present study, but 61% in the Jenkins et al. study). Hence, the controls in the Jenkins et al. study are relatives of the cases and thus, it is likely that they share parts of the polygenic background and other risk factors of their affected relatives (cases) to a certain extent which may explain the observed missing effect of the PRS. The comparison between population-based and registry-based predictions indicates that the study design and recruitment strategy may strongly influence the results and conclusions. Consequently, the application of PRS in clinical practice should consider the familial background and ascertainment of the patient.

Our data analyses provide evidence that the PRS acts as a relevant risk modifier for CRC among both the general population and population-based PV carriers in genes causing hereditary CRC. The findings of us and others qualify the PRS as important component of risk stratification and resulting risk-adapted surveillance strategies in terms of age of onset and frequency. Given the risk distribution across PRS groups, the PRS can define a considerable proportion of the general population at a CRC risk level which is considered sufficient for a more or a less intensive surveillance. Importantly, the non-carriers with high PRS are a much larger target group compared to PV carriers and thus might generate an even higher preventive effect form a healthcare perspective. A small group of non-carriers with positive FH and high PRS even has CRC risks almost in the same order of magnitude as LS carriers without additional risk factors and thus may need similar intensive surveillance measures.

According to these findings, there should be a potential benefit for both the general population and at-risk individuals carrying PV, from the inclusion of PRS in healthcare prevention policies, as risk-stratified surveillance improves early disease detection and prevention. A recent study demonstrated that individuals with a higher genetic risk benefited more substantially from preventive measures than those with a lower risk: CRC screening was associated with a significantly reduced CRC incidence and more than 30% reduced mortality among individuals with a high PRS.^49,50^ Preliminary calculations indicate that polygenic-risk-stratified CRC screening could become cost-effective under certain conditions including an AUC value above 0.65 which was reached in our analyses.^51^

Based on the striking different penetrance between individual hereditary CRC genes, very recent guidelines start to recommend a more gene-specific surveillance intensity in LS and polyposis.^52,53^ Given the strong modifying effect, the inclusion of additional risk factors will result in a more appropriate, clinically relevant risk stratification. Our results demonstrate that a combined risk assessment including FH and PRS will likely improve precise risk estimations and tailored preventive measures not only in the general population, but also in patients with hereditary disease.

Our study has some limitations. Firstly, there is evidence of a “healthy volunteers” selection bias of the UKBB population (UKBB participants tend to be healthier than the general population), and thus the results might not be completely generalizable in terms of effect sizes.^54^ Secondly, we cannot exclude that few carriers of APC PV who were classified as controls, are affected by a polyposis but have not been recognized as such or did not develop CRC due to intensive surveillance and / or prophylactic surgery, so that the calculated CRC risk of *APC* PV might be slightly underestimated. Thirdly, our risk assessment was based solely on genetic variants and FH and did not include other risk factors. Previous studies on UKBB showed that lifestyle modifiable risk factors play a pivotal role in cancer prevalence, and a shared lifestyle within families could influence FH with the disease.^47,55^ That might explain the partly independent association of the FH and the genetic risk. Finally, although we performed the analysis on the whole UKBB cohort, we could not test the risk stratification generalizability across different populations due to the limited sample size. PRS could be biased towards the European population as PRS was constructed based on European reference GWAS. Thus, these PRS might be a worse predictor in non-European or admixed individuals, as previously discussed in different studies.^56^

In conclusion, we show the important role of PRS and FH on CRC risk in both the general population and population-based carriers of a monogenic predisposition for CRC. The combined effect of common variants can strongly alter the age-related penetrance and life time risk of CRC. Thus, the PRS represents an additional, independent stratification level to cancer risk besides the FH and likely increase the accuracy of risk estimation. Consequently, PRS can define a relevant proportion within the general population as a risk group, which should be considered as subjects for more intense surveillance measures, and in addition point to a striking risk variability even among carriers of hereditary CRC, which requires more personalized, risk-adapted surveillance strategies. As expected, the modifying effect of the PRS seems to be relevant in particular for moderate penetrant CRC risk genes. When important factors such as polygenic background, FH, and non-genetic modifiers are included in risk assessment, the dichotomous risk division between sporadic and hereditary CRC will be partly replaced by a more continuous risk distribution.

## Data Availability

Genome-wide genotyping data, exome-sequencing data, and phenotypic data from the UK Biobank are available upon successful project application.
https://www.ukbiobank.ac.uk/

## Acknowledgements

UK Biobank analyses were conducted via application 52446 using a protocol approved by the Partners HealthCare Institutional Review Board. CM and EH are supported by the BONFOR-program of the Medical Faculty, University of Bonn (O-147.0002). This study was supported (not financially) by the European Reference Network on Genetic Tumour Risk Syndromes (ERN GENTURIS) – Project ID No 739547. ERN GENTURIS is partly co-funded by the European Union within the framework of the Third Health Programme “ERN-2016 – Framework Partnership Agreement 2017–2021”. DRB and PM are supported by the FNR INTER INTER/DFG/21/16394868. This research was also supported by the Instituto de Salud Carlos III and co-funded by European Social Fund—ESF investing in your future—(grants CM19/00099 and PID2019-111254RB-I00) and from the European Union’s Horizon 2020 research and innovation program under the EJP RD COFUND-EJP Nº 825575.

