## Supplementary material for "Clinically relevant combined effect of polygenic background, rare pathogenic germline variants, and family history on colorectal cancer incidence"

### Supplemental Figures

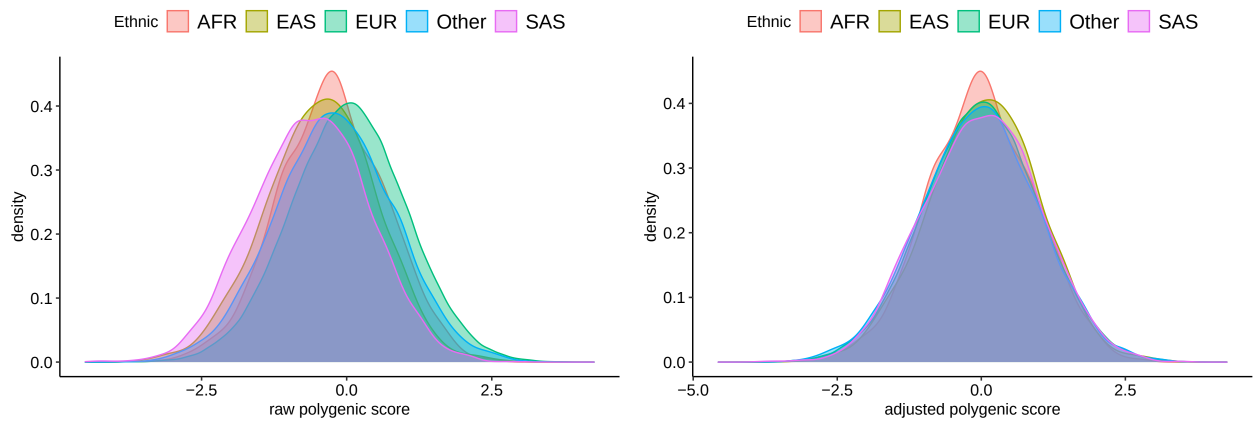


**(a)** raw PRS **(b)** adjusted PRS

**Figure S1:** Distributions of the colorectal cancer (CRC) PRS across the estimated genetic ethnicities. Estimated genetic ethnicities were estimated by projecting the samples in the 1000 genome project (1KGP) principal component space while considering the five 1KGP superpopulations as reference (https://github.com/privefl/paper-ancestry-matching/tree/master/code). Distributions of: a) raw PRS; b) adjusted PRS.

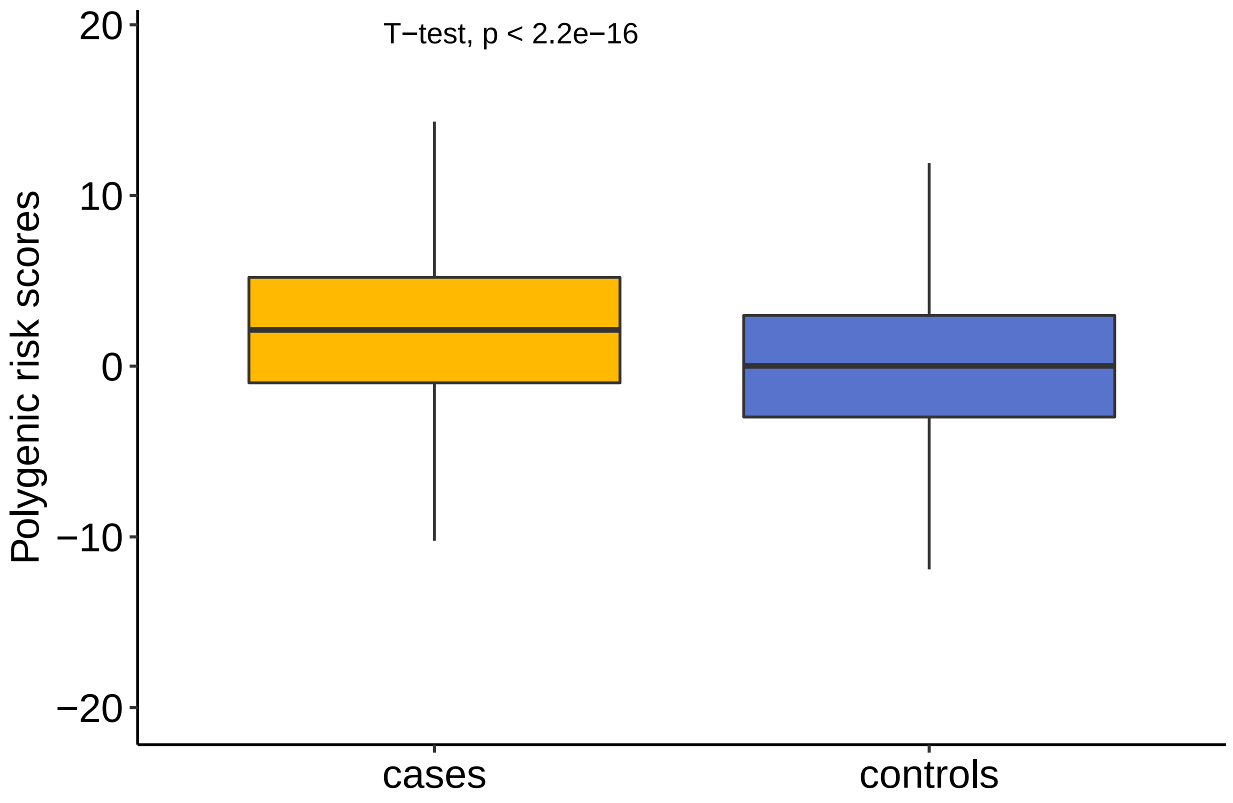


**Figure S2:** Adjusted PRS (aPRS) among CRC cases versus controls.

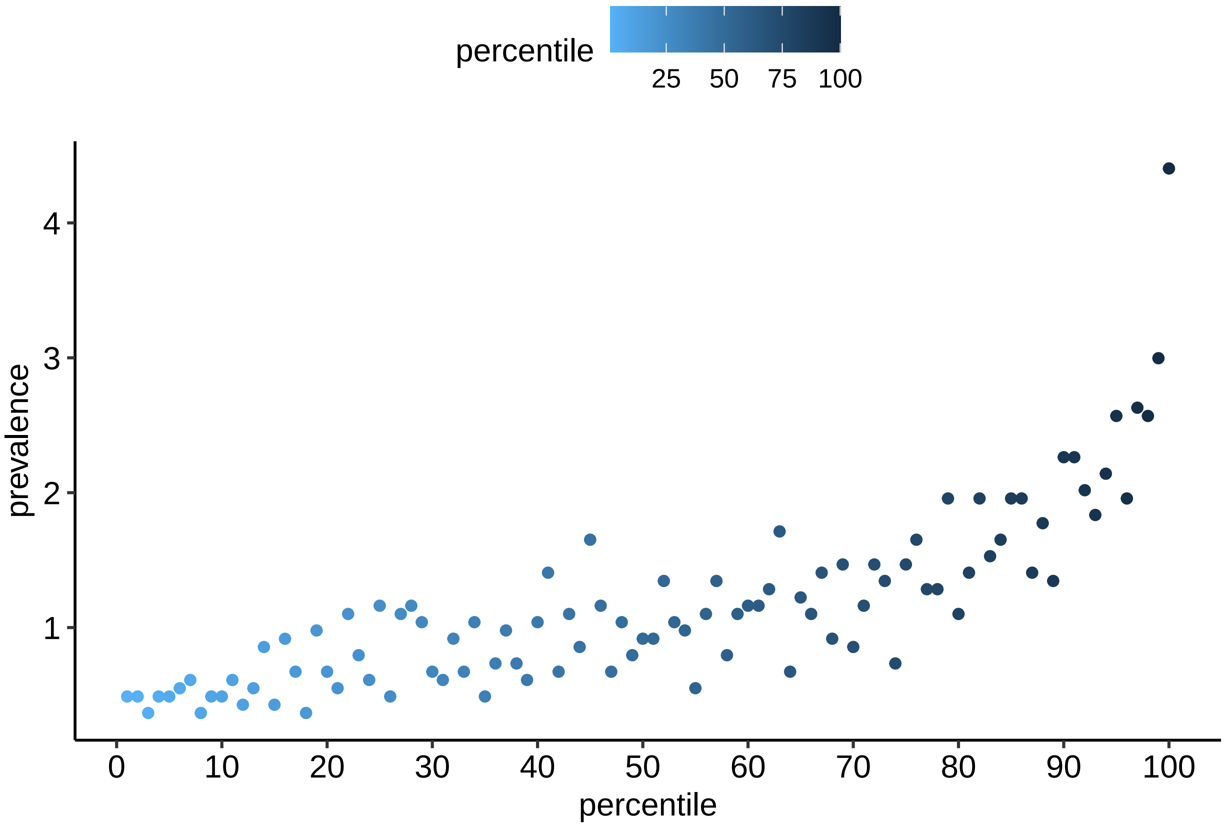


**Figure S3:** : Prevalence of the colorectal cancer (CRC) according to polygenic risk score (PRS) perecentiles.

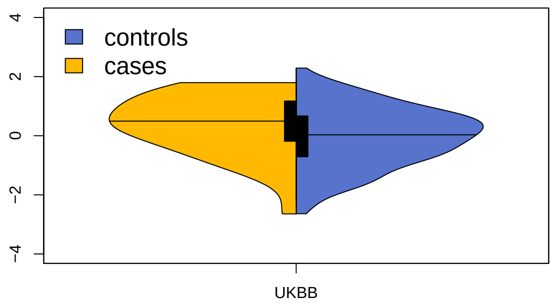


**(a)** Density plot

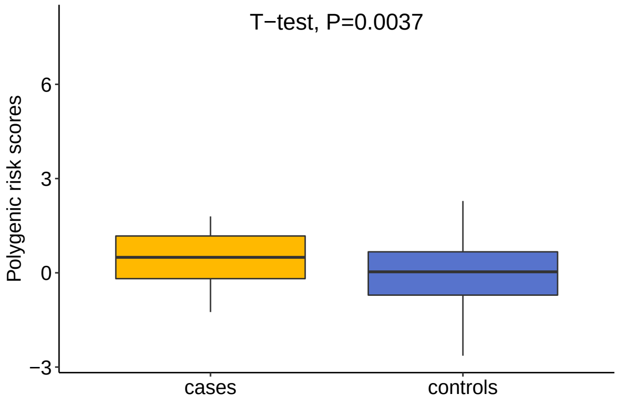


**(b)** Box plot

**Figure S4:** PRS among CRC affected (cases) and unaffected (controls) carriers. Density (a) and box plots (b) show the PRS distribution among affected and unaffected carriers. Horizontal line indicates PRS mean in plot (a).

### Supplemental Tables

**Table S1**: Provided as excel file.

**Table S2:** Number of PV carriers separated by gene and clinical status.

| gene | Cases | Controls | Total |
|---|---|---|---|
| APC | 3 | 8 | 11 |
| MLH1 | 6 | 5 | 11 |
| MSH2 | 2 | 11 | 13 |
| MSH6 | 14 | 121 | 135 |
| PMS2 | 5 | 224 | 229 |
| **Total** | **30** | **369** | **399** |
| MUTYH* | 5 | 365 | 370 |

*Note:*
* MUTYH variants were included only in the single gene analysis to compare the effect size with the other genes.

**Table S3:** Interplay of PV, FH, and PRS in CRC.

| | | Carrier | | | nonCarrier | | |
|---|---|---|---|---|---|---|---|
| prs | history | CI | OR | p.value | CI | OR | p.value |
| High | History | 97.8 | 39.9 (12.69-125.41) | 3e-10 | 25.53 | 3.08 (2.55-3.71) | <2e-16 |
| High | nonHistory | 62.1 | 14.56 (6.48-32.73) | 9e-11 | 21.03 | 2.16 (1.94-2.41) | <2e-16 |
| Intermediate | History | 66.4 | 15.63 (6.95-35.17) | 3e-11 | 16.29 | 1.83 (1.56-2.14) | 4e-14 |
| Intermediate | nonHistory | 42.5 | 5.56 (2.82-10.94) | 7e-07 | 10.37 | 1 | |
| Low | History | 50.1 | 5.78 (0.76-43.95) | 0.09 | 7.63 | 0.83 (0.56-1.24) | 0.4 |
| Low | nonHistory | 35.5 | 3.74 (0.91-15.43) | 0.07 | 6.16 | 0.57 (0.48-0.68) | 1e-10 |

*Note:*
CI = Cumulative incidence; OR = Odds ratio.
